# Does the timing of government COVID-19 policy interventions matter? Policy analysis of an original database

**DOI:** 10.1101/2020.11.13.20194761

**Authors:** M. Stephens, J. Berengueres, S. Venkatapuram, I. A. Moonesar

## Abstract

**Objective:** Though the speed of policy interventions is critical in responding to a fast spreading pandemic, there is little research on this topic. This study aims to (1) review the state of research on the topic (2) compile an original dataset of 87 COVID-19 non-pharmaceutical interventions across 17 countries and (3) analyses the timing of COVID-19 policy interventions on mortality rates of individual countries.

**Design:** Statistical analysis using Excel and R language version 3.4.2 (2017-09-28) of 1479 non-pharmaceutical policy interventions data points.

**Setting:** China, Singapore, South Korea, Japan, Australia, Germany, Canada, India, United Arab Emirates, United States of America, South Africa, Egypt, Jordan, France, Iran, United Kingdom and Italy.

**Population:** 36 health policies, 19 fiscal policies; 8 innovation policies; 19 social distancing policies, and 5 travel policies – related to COVID-19.

**Interventions:** We calculate the time (time-lag) between the start date of a policy and three-time specific events: the first reported case in Wuhan, China; the first nationally reported disease case; the first nationally reported death.

**Main Outcome Measures:** National level mortality rates across 17 countries. Mortality rate is equivalent to (death attributed to COVID-19) / (death attributed to COVID-19 + COVID-19 recovered cases).

**Results:** The literature review found 22 studies that looked at policy and timing with respect to mortality rates. Only four were multicountry, multi-policy studies. Based on the analysis of the database, we find no significant direction of the association (positive or negative) between the time lag from the three specified points and mortality rates. The standard deviation (SD) of policy lags was of the same order of magnitude as the mean of lags (30.57 and 30.22 respectively), indicating that there is no consensus among countries on the optimal time lags to implement a given policy. At the country level, the average time lag to implement a policy decreased the longer the time duration between the country’s first case and the Wuhan first case, indicating countries got faster to implement policies as more time passed.

**Conclusions:** The timing of policy interventions across countries relative to the first Wuhan case, first national disease case, or first national death, is not found to be correlated with mortality. No correlation between country quickness of policy intervention and country mortality was found. Countries became quicker in implementing policies as time passed. However, no correlation between country quickness of policy intervention and country mortality was found. Policy interventions across countries relative to the first recorded case in each country, is not found to be correlated with mortality for 86 of the 87 policies. At the country level we find that no correlation was found between country-average delays in implementing policies and country mortality. Further there is no correlation with higher country rankings in The Global Health Security Index and policy timing and mortality rates.

**Funding Statement:** This work was supported by the Alliance for Health Policy and Systems Research at the World Health Organization as part of the Knowledge to Policy (K2P) Center Mentorship Program.

**A competing interests statement”:** All authors have completed the ICMJE uniform disclosure form at www.icmje.org/coidisclosure.pdf and declare: IAM and MS would like to acknowledge the Alliance for Health Policy and Systems Research at the World Health Organization for financial support for publishing as part of the Knowledge to Policy (K2P) Center Mentorship Program [BIRD Project].

## INTRODUCTION

With regard to policymaking during a pandemic, there are several assumptions being made. First, there is an assumption being made that early policy interventions would lower transmission rates and mortality rates.^1 2 3^ However, this generally accepted view of the relevance and importance of timing of policy interventions from past epidemics is supported by very few studies. ^4 5 6^ The World Health Organization Writing Group stresses that pandemic interventions are often based on limited information and vary depending on the context.^7^ There are competing concerns that countries need to manage, and these may be influenced by internal events (elections) or external events (global reputation).^8 9^

Often the urgency of policy interventions focuses on one of two effects – saving lives and staunching the effects of the disease on the economy.^10 11 12^ Existing studies of COVID-19 policy interventions look at policies in isolation like travel restrictions,^13^ contact tracing,^14^ proactive testing,^15^ isolation, and social distancing,^16^ lockdowns,^17 18^ or school closure.^19^ Single policy studies may not accurately reflect the complexity of the situation (health, economy, societal concerns, for example). The World Health Organization recommends whole-of-society coordination mechanisms to support preparedness and response, including the health, transport, travel, trade, finance, security and other sectors”.^20^ A recent study on the timing for social distancing, in Wuhan (Hubei, China), South Korea, Japan, Hong Kong, Singapore, and Italy between January and March 2020 finds that the implementation of the social distancing policy was random.^21^ There are few multi-policy studies available.

In a majority of cases,where those studies exist, they examine policy implementation in a single country and assume the findings are transferable across borders.^22 23^ Few multi-country, multi-policy studies exist. We found one study^24^ that looked at 17 policies, but all focused on various types of social distancing measures. The countries studies were China (Wuhan), South Korea, Japan, Hong Kong, Singapore, and Italy. The study finds that the information available was sparse and based on what they collected, they state that it is difficult to quantitatively assess the efficacy of many interventions, though they do conclude that social distancing slows the spread of the disease. On the other hand, a muti-country, multi-policy study covering China, South Korea, Italy, Iran, France, and the United States, focusing on travel restrictions, social distancing through cancellations of events, suspensions of educational/commercial/religious activities, quarantines, lockdowns, state of emergency declarations, and expansions of paid sick leave, found that policies slow COVID-19 virus contagion as measured by cases and deaths.^25^

Some studies have cautioned against applying learnings from previous pandemics to the present one, and stress that more data is needed on interventions and their impact in individual countries.^26 27^ Due to the relatively little research across countries and the robustness of findings on the timing of policy interventions and mortality, this study aims to evaluate the timing of policy intervention and their correlation to mortality rates. This research will (1) conduct a systematic review of COVID-19 papers that study the impact of the timing of the policies intervention on mortality rates (2) compile a dataset of COVID-19 policy interventions across 17 countries (3) evaluate the correlation of the quickness of a country-level policy intervention on country mortality rate and (4) evaluate given policy across countries, to assess if there is a correlation between quickness and mortality rate.

## METHODS

We conducted a systematic review of studies on the impact of timing of policy intervention on mortality rates during the COVID-19 period. The search was limited to COVID-19 policies and interventions published in the English language for articles indexed in PubMed and Proquest that were published between January 2020 and May 2020. The keywords included: COVID-19 ‘timing of policy’, ‘non-pharmaceutical interventions, epidemics/pandemics, health crisis, outbreaks, on mortality rates.

To create the dataset, we identified 87 COVID-19 policy interventions across 17 countries, daily COVID-19 cases, and mortality rates. The start date is from December 31 2019, and the end date is May 16 2020. The data collection was conducted between April 6 2020, and June 4 2020. The start dates of 87 policy interventions were collected using various sources such as OxCGRT,^28^ data from Institute Montaigne, government websites, research papers, press releases, press conferences, and newspaper reports. In many cases, the start date of the policy is not explicitly given. We use the date of the first announcement of the policy. Once the start date was determined, the lag in days between the start date and (1) the first case in Wuhan (2) first case in the country and (3) the first confirmed death in the given country was then calculated. The assumption we make is that the policy is deployed in response to the local COVID-19 situation.^29^

Deaths and cases were compiled using daily reports from the European Centre for Disease Prevention and Control. We decided to use the mortality rate to compare countries as it is a more robust measure of the pandemic outcomes. We define mortality rate as (death attributed to COVID-19) / (deaths + recovered cases).^30^

The 17 countries chosen for analysis are among the first affected countries and are identified as having medium to high rankings on the 2019 Global Health Security Index (see Table 1). Three critical events happened during the time the sample countries had their first infection: Wuhan Lockdown (3 countries), Chinese New Year (which was a significant milestone for transmission of the disease)^31^ and WHO declaration of Public Health Emergency (6 countries), and WHO declaration of pandemic (8 countries). All these events could be considered as early warning signals for governments to plan policy interventions (Appendix 1). The countries are representative of three continents Asia, Europe, and North America.

**Table 1:**
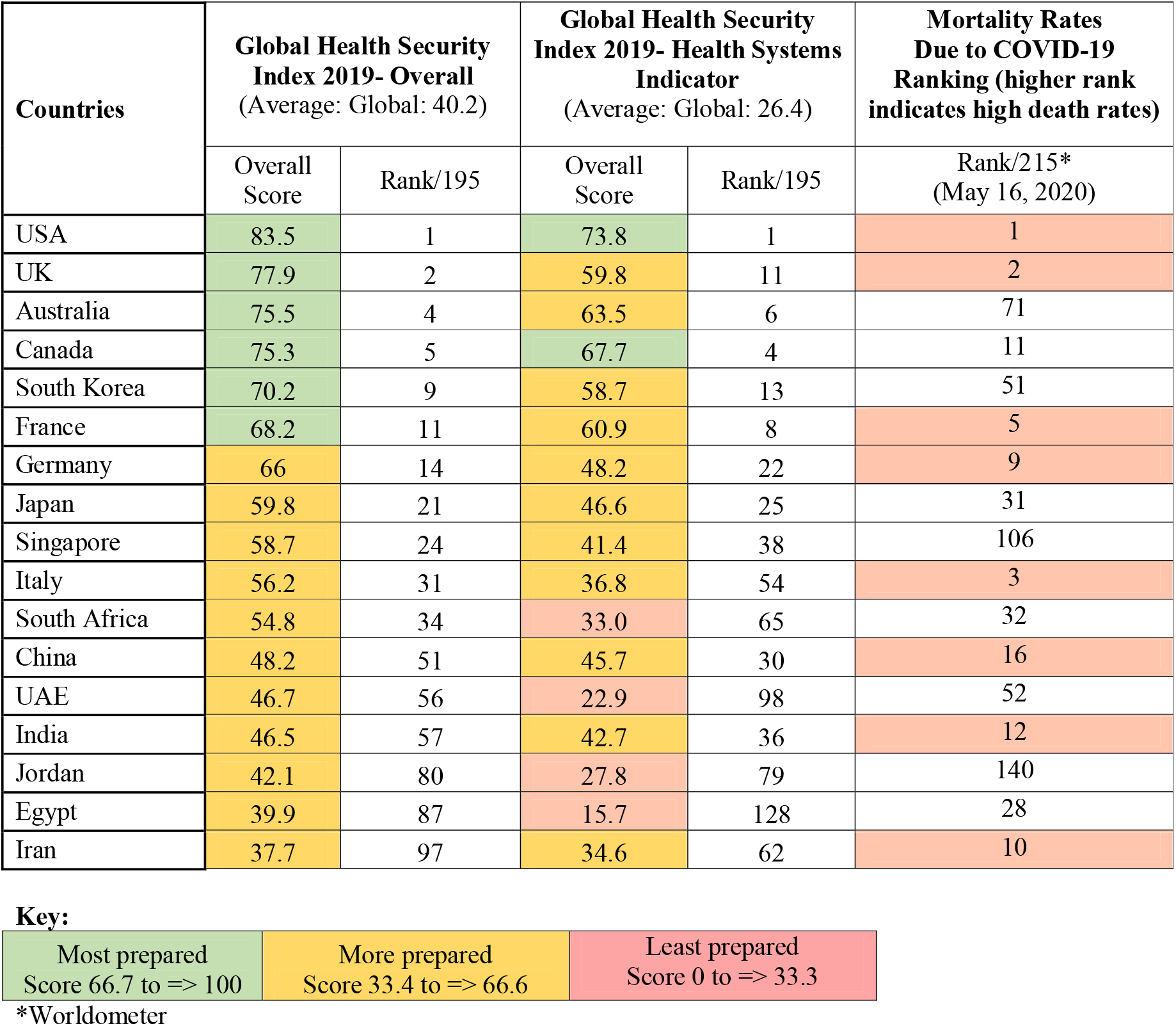
Ranking of Sample Countries

| Countries | Global Health Security Index 2019- Overall<br>(Average: Global: 40.2) |  | Global Health Security Index 2019- Health Systems Indicator<br>(Average: Global: 26.4) |  | Mortality Rates Due to COVID-19 Ranking (higher rank indicates high death rates) |
| --- | --- | --- | --- | --- | --- |
|  | Overall Score | Rank/195 | Overall Score | Rank/195 | Rank/215*<br>(May 16, 2020) |
| USA | 83.5 | 1 | 73.8 | 1 | 1 |
| UK | 77.9 | 2 | 59.8 | 11 | 2 |
| Australia | 75.5 | 4 | 63.5 | 6 | 71 |
| Canada | 75.3 | 5 | 67.7 | 4 | 11 |
| South Korea | 70.2 | 9 | 58.7 | 13 | 51 |
| France | 68.2 | 11 | 60.9 | 8 | 5 |
| Germany | 66 | 14 | 48.2 | 22 | 9 |
| Japan | 59.8 | 21 | 46.6 | 25 | 31 |
| Singapore | 58.7 | 24 | 41.4 | 38 | 106 |
| Italy | 56.2 | 31 | 36.8 | 54 | 3 |
| South Africa | 54.8 | 34 | 33.0 | 65 | 32 |
| China | 48.2 | 51 | 45.7 | 30 | 16 |
| UAE | 46.7 | 56 | 22.9 | 98 | 52 |
| India | 46.5 | 57 | 42.7 | 36 | 12 |
| Jordan | 42.1 | 80 | 27.8 | 79 | 140 |
| Egypt | 39.9 | 87 | 15.7 | 128 | 28 |
| Iran | 37.7 | 97 | 34.6 | 62 | 10 |

**Key:**
|  |  |  |
| --- | --- | --- |
| Most prepared<br>Score 66.7 to => 100 | More prepared<br>Score 33.4 to => 66.6 | Least prepared<br>Score 0 to => 33.3 |

\*Worldometer

Table 1 presents a ranking of countries ranked as ‘most prepared’, ‘more prepared’, and ‘least prepared’ from the Global Health Security Index 2019^32^ and the Worldometer COVID-19 Mortality Ranking as of May 16, 2020.^33^

## ANALYSIS

The review identified 142 relevant studies focusing on COVID-19 related non-pharmaceutical intervention (NPI) policies. After screening by abstract/title, 28 studies were selected for full-text assessment. Of them, 22 were finally selected (see Appendix 2). The majority of the studies focused on a single country (n=18) or a small group of countries (n=4) with similar policy interventions (for example, school closure (n=8). China was the most studied country (37%, n=8), followed by the USA (27%, n=6), then Iran (1), Canada (1), Lebanon (1), and Taiwan (1). There were four multiple country studies: West Africa as a region, a global study on YouTube videos with no specific country identified; one looking at China, Iran, Japan, Italy, and South Korea; and a two-country study looking at Spain and the UK. The study period for these studies was from December 2019 to May 2020. The majority of studies used a quantitative research design (n=19), where the main focus was on timely and accurate dissemination of policies in terms of managing during pandemics.

Though these studies used the word timing, they did not investigate the impact of the timing of the policy intervention on mortality rates during the COVID-19 period. Instead, these studies looked at which public policy measures and interventions were critical in limiting the spread of COVID-19 over a particular time period.^34 35 36 37^ One study suggested that policymakers could have the best chance for policy learning due to the time lag between the China outbreak COVID-19 and the spread in the context of Iran.^38^ The papers have highlighted the need for the importance of delivering timely and accurate information.^39 40 41 42 43^

Our original dataset analysis was at two levels: looking at time lags at the country-level and the policy-level. We treat time lags in a similar way, as previously done in a big data modeling application.^44^ The time lag between policy implementation and a COVID-19 milestone was defined as the number of days between the policy start date and the given milestone date. We run statistical models for three different milestones: (i) the first case confirmed in the country, (ii) the first death in the country and, (iii) the date of the first case in Wuhan, China, and their correlation to COVID-19 mortality rates. The effectiveness and degree of enforcement have not been evaluated here, as there can be variance in implementation across geographical regions within a country, particularly in large federal governance structures, autonomous regions, and so forth. Hence, each policy time lag is relative to one global event (China first case) and two country-specific milestones.

At the policy level, to assess the linear association and direction between country mortality and policy time lag, we used the Spearman’s rank correlation coefficient. However, both Spearman and Pearson yielded similar results for the three different milestones considered. Table 2 shows a matrix of time lags (each cell is a time lag) where rows are the 87 policies and columns are the 17 countries. Each cell indicates the distance in days between the first confirmed case and the start of the policy in each given country. A negative figure indicates how many days before the first confirmed case, the policy was implemented. A positive number shows how many days the policy was implemented *after* the first confirmed case. A blank indicates that there is no data available because the policy was either never implemented in the given country or information is not available.

**Table 2.**
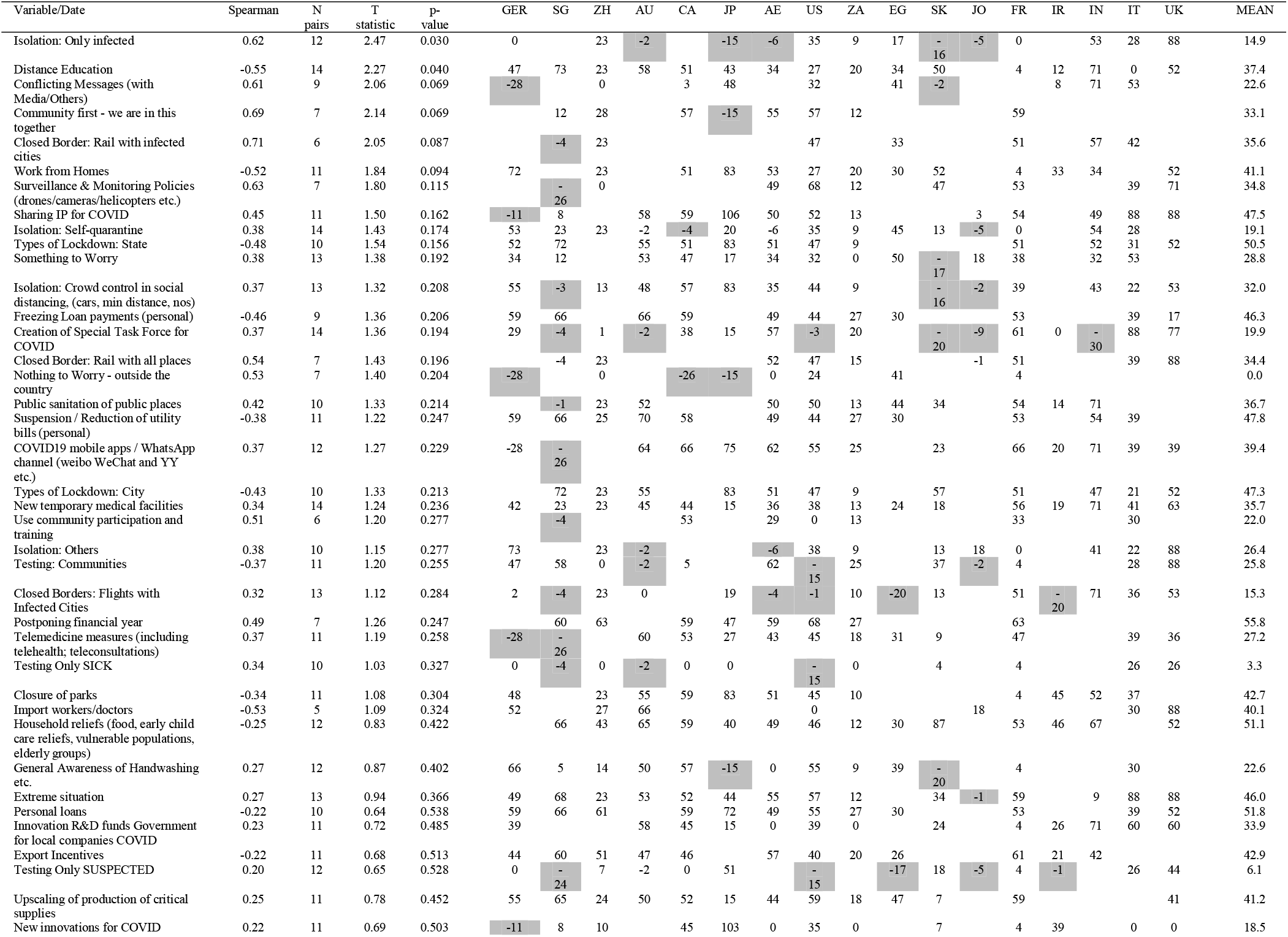
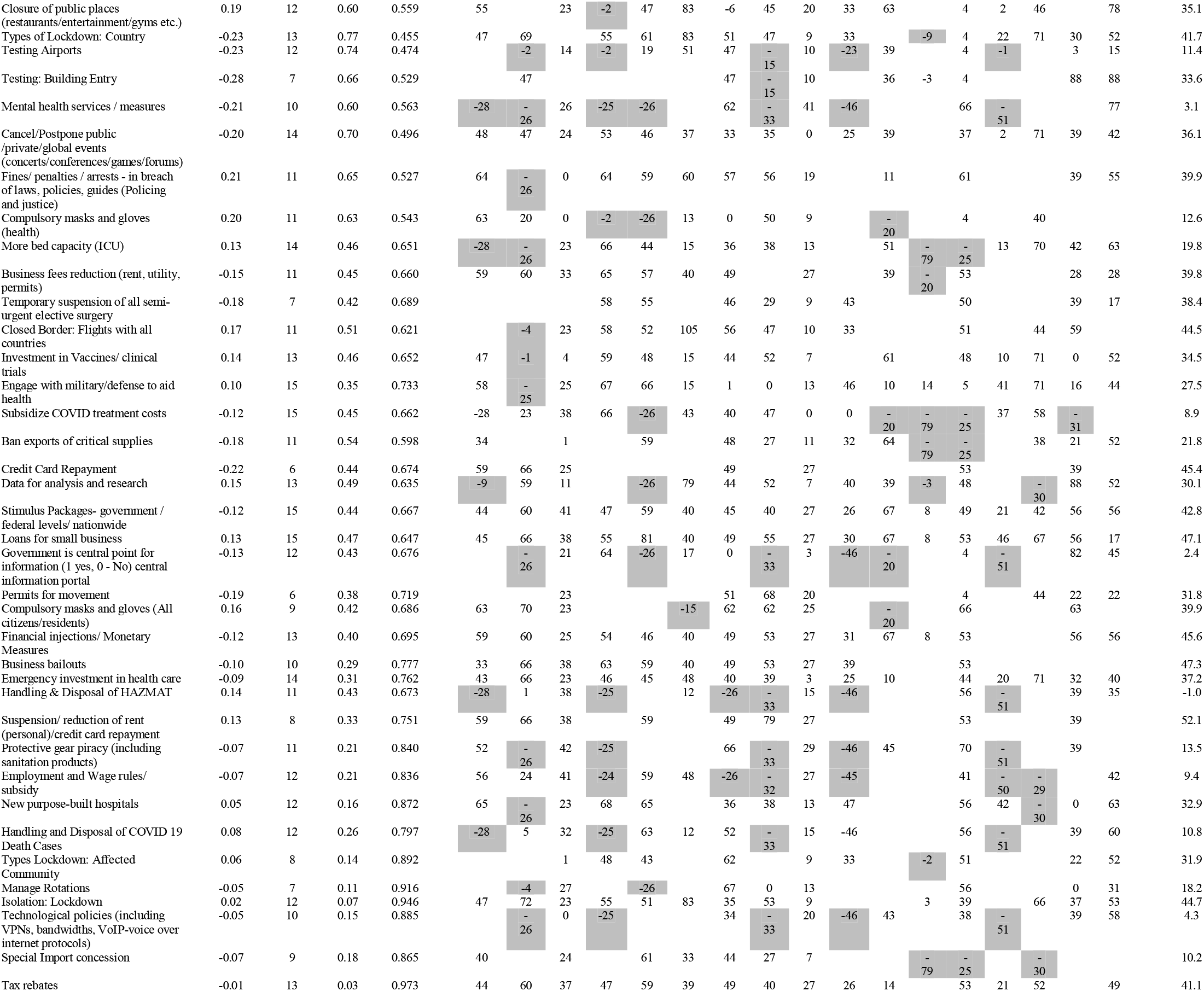
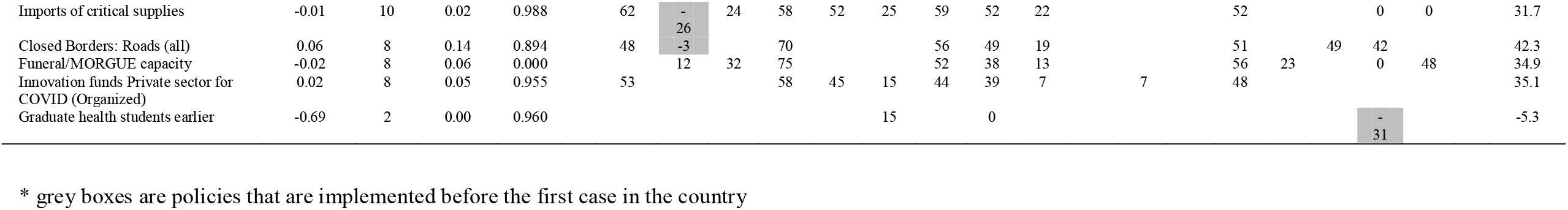
List of policies and their implementation delay (time delay/time lag variable) since the first confirmed case for each country.

For each policy, we calculate the correlation with country-specific indicators such as mortality rate, which is equivalent to (death attributed to COVID-19) / (death attributed to COVID-19 + COVID-19 recovered cases). We choose this indicator because it reflects how effectively the country is coping. Then we compute the Spearman rank correlation coefficient, confidence intervals, and p-values. For reference, we also computed Pearson coefficients. The same analysis, mentioned above, was conducted with different COVID-19 curve milestones (N^th^ death, M^th^ case). The results are similar and provided in the appendix. A dynamic Excel sheet is also provided where N and M are parameters that can be modified, and the Pearson and Spearman coefficients are re-calculated on the go. The analysis was conducted both in Excel and R language (version 3.4.2 2017-09-28). The Spearman p-values were computed both in Excel and in R using the SpearmanCI package. Slight differences (+/-5%) in the Spearman coefficient values due to different slight formulas used by both Excel and SpearmanCI are also noted, and confirm the differences stated previously by the authors of the SpearmanCI package.^45^ For 86 policies, no significant correlation was found. Only one policy was found to be weakly correlated, the policy for isolation of the infected, with a sample size of 12 countries applying the policy, and a Spearman Coefficient =0.63; (95% CI [0.27 - 0.91] p-value 0.03). See Figure 1.

**Figure 1:**
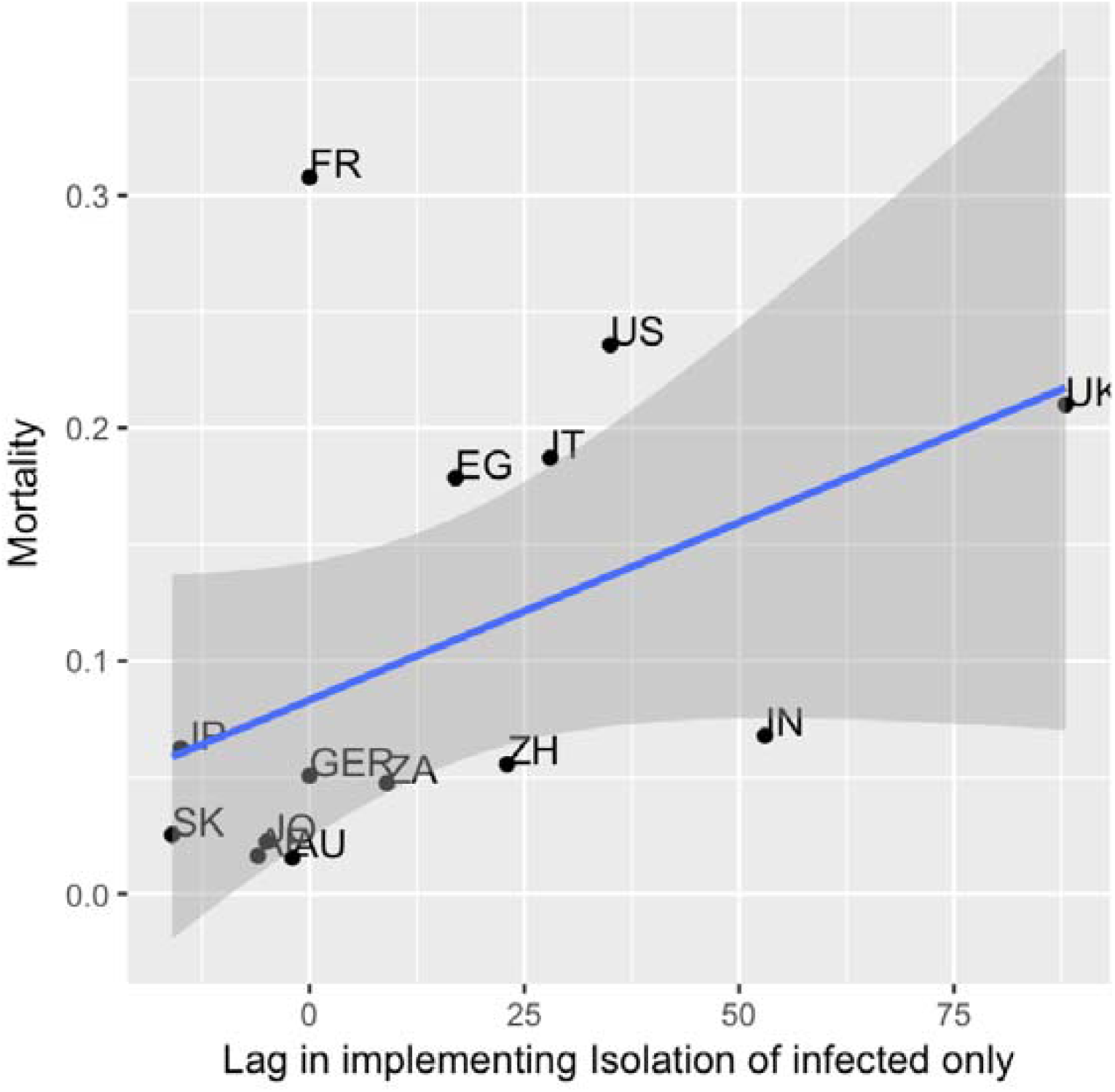
Lag in implementing Isolation policy at the Country Level

At the country level, we ran a linear regression to quantify whether countries implement policies quicker as time lapsed (see Table 3 and Figure 2). In the regression, the days elapsed since each country’s reported first case and the Wuhan first case was the dependent variable. The independent variable is the mean of each policy time lags for each country. The linear regression yields Multiple R-squared: 0.511, Adjusted R-squared: 0.476, and a p-value: 0.001. Intercept 45.51, slope – 0.55. For every day elapsed since the Wuhan first case was reported, countries reduced the average delay to implement a policy relative to their own first case by half-day approximately. However, no correlation was found between country-average delays in implementing policies and country mortality. Furthermore, no conclusive association was found either between the Global Health Security Index scores of countries and mortality (Figure 3).

**Table 3.** Country-level statistics and stats on time lags since the first confirmed case.

|  | GER | SG | ZH | AU | CA | JP | AE | US | ZA | EG | SK | JO | FR | IR | IN | IT | UK |
| --- | --- | --- | --- | --- | --- | --- | --- | --- | --- | --- | --- | --- | --- | --- | --- | --- | --- |
| Rank of Mortality | 7 | 1 | 8 | 2 | 12 | 9 | 3 | 15 | 6 | 13 | 5 | 4 | 16 | 10 | 11 | 14 | 15 |
| Mortality(D/D+R) | 0.05 | 0.01 | 0.06 | 0.02 | 0.13 | 0.06 | 0.02 | 0.24 | 0.05 | 0.18 | 0.03 | 0.02 | 0.31 | 0.06 | 0.07 | 0.19 | 0.21 |
| Max - Min | 101 | 99 | 63 | 100 | 107 | 121 | 93 | 112 | 41 | 96 | 107 | 97 | 95 | 97 | 101 | 119 | 88 |
| Max | 73 | 73 | 63 | 75 | 81 | 106 | 67 | 79 | 41 | 50 | 87 | 18 | 70 | 46 | 71 | 88 | 88 |
| Min | -28 | -26 | 0 | -25 | -26 | -15 | -26 | -33 | 0 | -46 | -20 | -79 | -25 | -51 | -30 | -31 | 0 |
| Count | 62 | 67 | 69 | 61 | 63 | 57 | 76 | 80 | 76 | 45 | 48 | 26 | 79 | 35 | 44 | 66 | 57 |
| Median | 47 | 12 | 23 | 54 | 52 | 40 | 49 | 39.5 | 13 | 30 | 20.5 | -2 | 51 | 14 | 52 | 39 | 52 |
| SD. S | 32.4 | 36.7 | 14.3 | 31.9 | 28.9 | 32.5 | 23.0 | 29.5 | 8.8 | 31.7 | 29.1 | 30.9 | 26.5 | 32.5 | 30.7 | 25.1 | 22.2 |
| Mean | 33.9 | 22.7 | 22.7 | 39.0 | 40.6 | 39.6 | 38.8 | 30.3 | 15.1 | 17.4 | 23.9 | -11.0 | 35.5 | 5.9 | 45.6 | 35.0 | 51.8 |
| Mth case | 1/28/2<br>0 | 1/26/2<br>0 | 12/31/1<br>9 | 1/25/2<br>0 | 1/26/2<br>0 | 1/15/2<br>0 | 1/29/2<br>0 | 2/2/20 | 3/6/20 | 2/15/2<br>0 | 1/20/2<br>0 | 3/19/2<br>0 | 1/25/2<br>0 | 2/20/2<br>0 | 1/30/2<br>0 | 1/31/2<br>0 | 1/7/20 |
| Nth death | 3/10/2<br>0 | 3/22/2<br>0 | 1/11/20 | 3/1/20 | 3/10/2<br>0 | 2/13/2<br>0 | 3/16/2<br>0 | 2/29/2<br>0 | 3/27/2<br>0 | 3/9/20 | 2/21/2<br>0 | 3/29/2<br>0 | 2/15/2<br>0 | 2/20/2<br>0 | 3/13/2<br>0 | 3/13/2<br>0 | 2/18/2<br>0 |
| M-N | 42 | 56 | 11 | 36 | 44 | 29 | 47 | 27 | 21 | 23 | 32 | 10 | 21 | 0 | 43 | 42 | 42 |
| Days elapsed since Wuhan<br>first death to country first<br>death | 59 | 71 | 0 | 50 | 59 | 33 | 65 | 49 | 76 | 58 | 41 | 78 | 35 | 40 | 62 | 62 | 38 |
| **Days elapsed since Wuhan**<br>**first case to country first case** | 28 | 26 | 0 | 25 | 26 | 15 | 29 | 33 | 66 | 46 | 20 | 79 | 25 | 51 | 30 | 31 | 7 |

\* D = death and R= recovered, M=1, N=1

**Figure 2:**
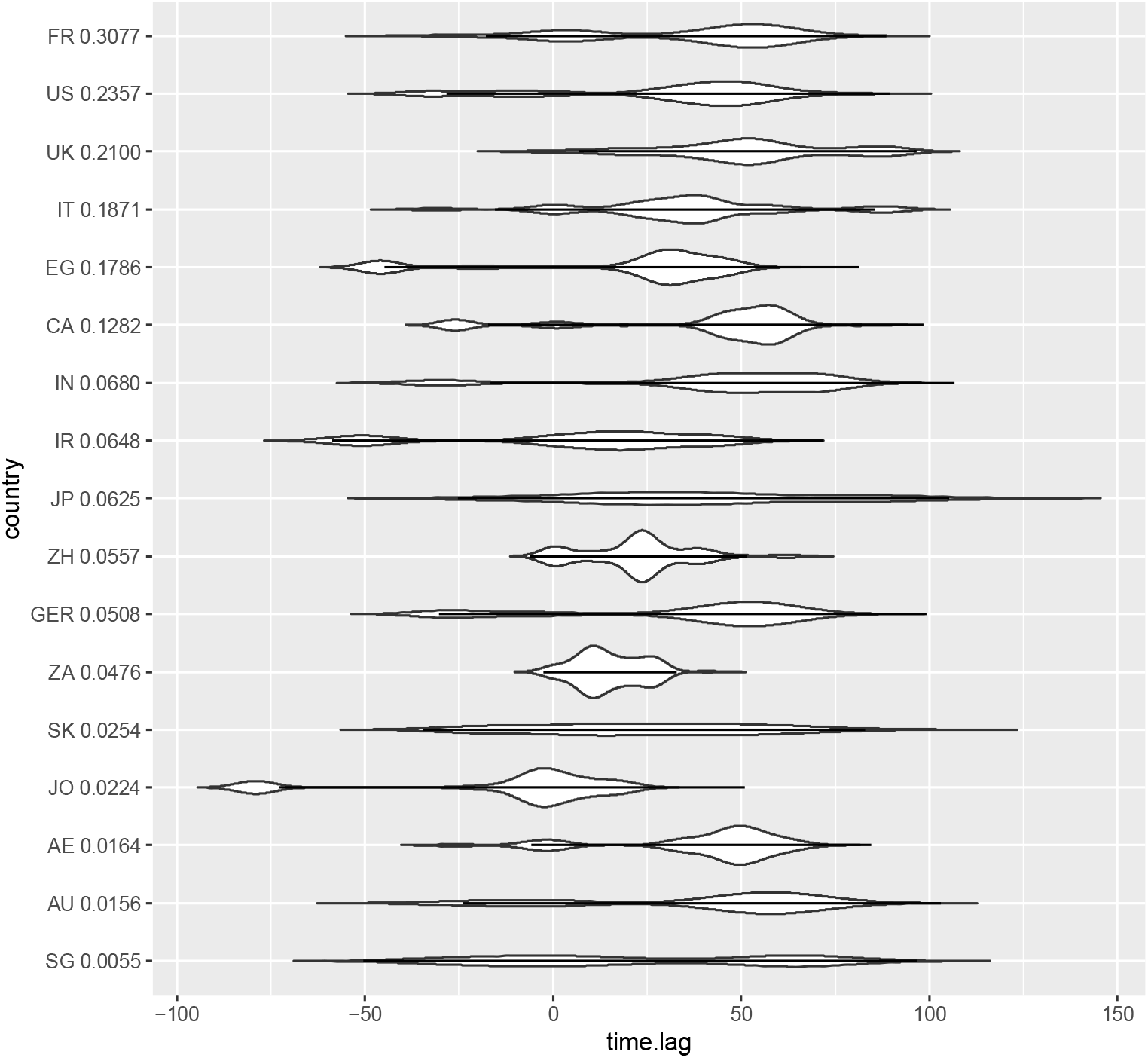
Density of new COVID-19 policies for the 17 countries considered. Countries ordered by descending mortality. Y is density of new COVID-19 policies. X is time lag since country first case.

**Figure 3:**
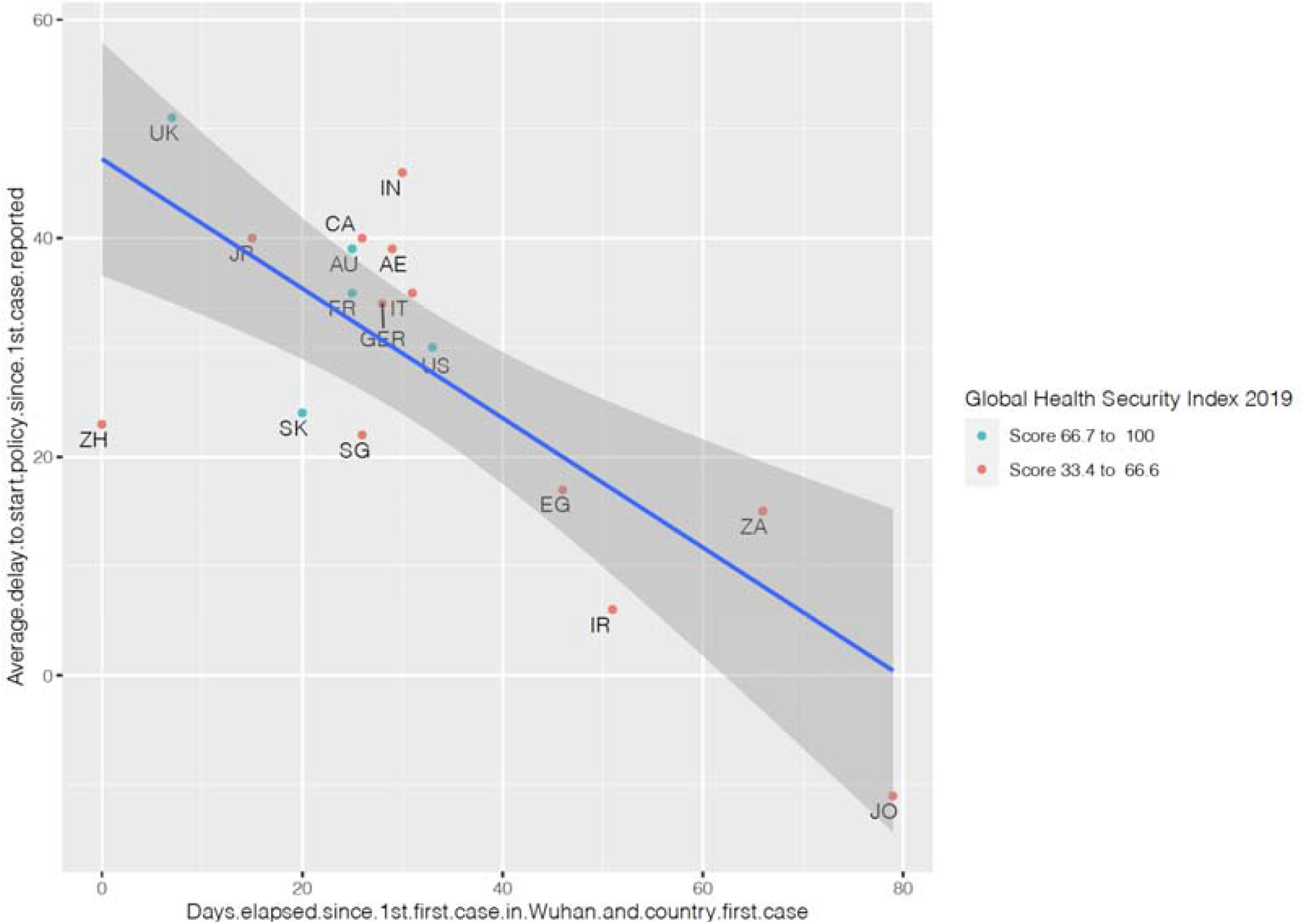
Country-average delay in implementing a policy and country mortality

We find that having a higher score in the 2019 Global Health Security Index did not mean the country handled the pandemic better (see Figure 3).

Figure 4 shows the distribution of time-lags relative to the first confirmed case in each country. Mortality rates are ranked by the countries (least to highest). This diagram highlights the fact that policy delays or policy implementation quickness are not correlated to mortality rate.

**Figure 4:**
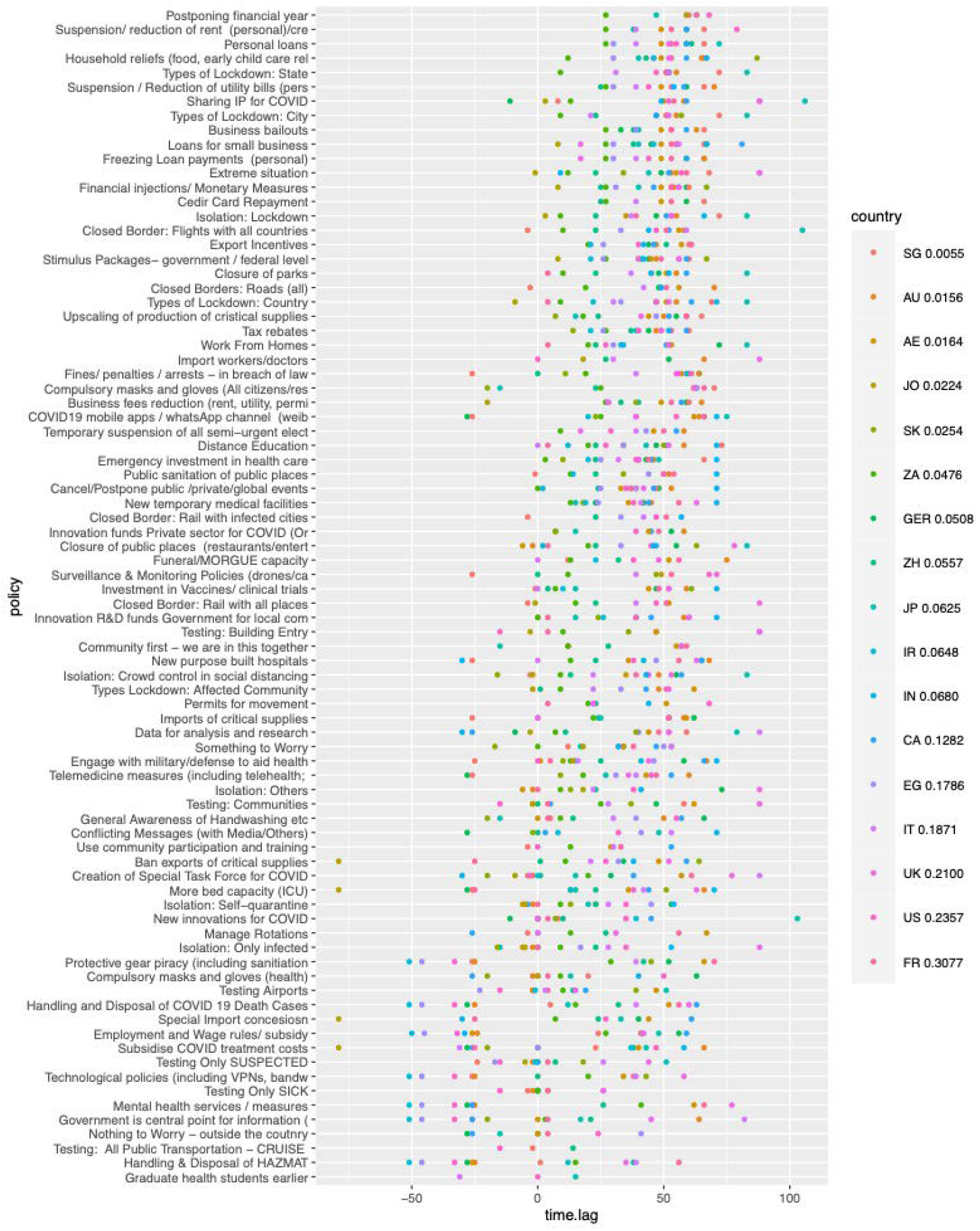
Time-lag per policy for each country

## FINDINGS

### No Correlation found between Policy Timing and Mortality Rate

We examined 87 policies across 17 countries. The timing of policy interventions in and across countries relative to (1) first case in Wuhan, China, (2) first recorded case in each country and (3) first recorded death in each country, is not found to be correlated with mortality, though we find that countries became quicker in introducing policy interventions as time passed. This finding supports the recent conclusions of other researchers^46^ who state, “*countries that have flattened death curves earliest may not provide a basis for extrapolating trends in areas where similar control could prove elusive*.” Interventions likely had reinforcing effects and were driven by critical behaviour changes within local communities^47^ and perhaps existing conditions in public health infrastructure.

No significant direction of the association was found (positive or negative) except for one policy (“Isolation of only infected”). This policy showed a positive association between the quickness to implement the policy and mortality (Spearman coefficient +0.61 95% CI [0.27 - 0.91]). Other researchers found that isolating all positive cases, including the asymptomatic, helped in containing the virus. ^48^

### Policies across countries are not implemented with the same urgency

The SD of policy time lags was of the same order of magnitude as the mean of lags (30.57 and 30.22 respectively), indicating that there is no agreement between countries on what are the optimal times to start a given policy (see Figure 5). This is surprising as a pandemic requires some coordination across borders. This was a fast-spreading virus. In our sample, three countries identified their first case from the first case in China to January, 6 countries between the Wuhan Lockdown and the Chinese New Year/WHO Global alert (8 days), and another 8 countries between the WHO Global Alert and Pandemic declaration (40 days).

**Figure 5:**
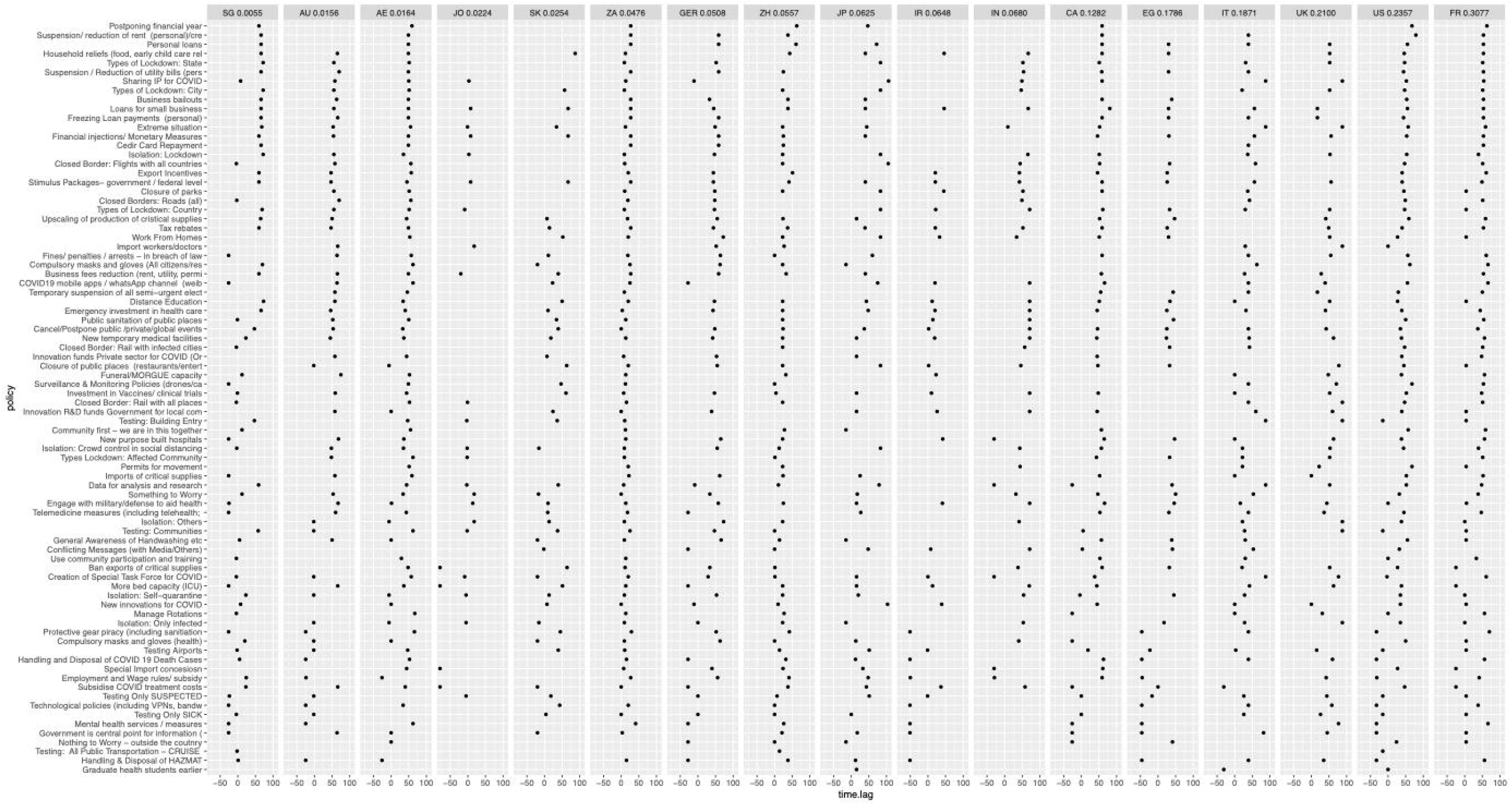
Time-lag of policy concerning the first case across countries

### Countries implement policies faster from Wuhan first case

The average time to implement a policy decreased the farther the country’s first case was dated from the Wuhan first case, indicating that countries became quicker in policy responses, which may also indicate that they also could have become more pre-emptive in policy responses as the pandemic evolved. This finding is in line with the Oxford policy study that finds internationally, government policy responses have become more stringent as more time passes.^49^

### No correlation between country ranking and country policy timing implementation

We find no correlation with higher country rankings in The Global Health Security Index and policy timing and mortality rates. Such indexes do not capture those “other” factors^50 51^ or help in identifying the factors that were instrumental in some countries managing the pandemic response better (geographical isolation, population conformity to mask-wearing). Such country-specific factors can span social,^52^ environmental, and political factors^53^, often requiring behavioral insights,^54^ not captured in many quantitative models.

## DISCUSSION

Our findings indicate there needs to be stronger global coordination across countries and coordination within countries (federal and local) on policy formulation, timing, and implementation. Our study suggests that the limited impact of policy timing on mortality may arise if the country does not have the underlying systems, strategies, and culture in place. While WHO does highlight the importance of multi-sector planning for non-pharmaceutical public health measures,^55^ the reports for guidance are outdated: the only existing pandemic report focuses only on influenza is dated 2009^56^; the other relevant reports on infection prevention and control (IPC) programmes is dated 2019 and the “Managing epidemics” is dated 2018.^57^

The quality of timely data that needs to be shared within countries and across borders needs to be increased. One of the challenges of studies like this is understanding the time it takes for a policy to be implemented and for it to take effect. A policy announcement does not automatically mean that a policy implementation took place. Two studies found that the announcement of the public health intervention and its subsequent implementation, showed little evidence of any impact.^58 59^ At times, the signal that the policy introduction communicates may provide an illusion of impact (like in monetary policies and stock market reactions).^60^

## CONCLUSION

Top-down policy implementation may not translate as relevant to a bottom-up perspective.^61^ This could be the issue of coordination, execution, or the political will not be resonating with communal interests.^62 63 64^ Policy implementation presupposes collaborative governance – where relevant stakeholders have an incentive or history of working together towards a common outcome.^65^ Policy failure can occur as a result of bad execution, bad policy, or bad luck.^66^

We suggest future research draw on a more extensive dataset of all country responses, the easing of COVID-19 policy responses, and study the reinforcing effects of policies on society. This is because socio-demographic heterogeneity and country policy differences across national borders, interventions may have different effects. Caution needs to be taken when extrapolating learnings across countries.^67^

## STRENGTHS OF THE STUDY

- This study was an exhaustive policy study that involved manually compiling a policy database of and 1479 policies and case and mortality data for the period between December 31 2019 to May 31 2020.
- Visualization of the results allow policy makers to have greater clarity on responsiveness to the pandemic.

## LIMITATIONS OF THE STUDY

- The term timing has been used fluidly in literature reviews and most studied do not focus on when the policy was introduced but before and after a policy intervention. In our case the word timing refers to quickness of response of policy intervention and its impact on mortality.
- Policy implementation dates are based on announcement as there is no clarity on level of implemntion.
- In some cases policy interventions are announced at a city, region or state-level however in that case we have still assumed it is the first date of implementation in that country.
- The findings may change as the sample size increases and as policy interventions ease.

## Supporting information

Literature Review

IAM COI Disclosure

JB COI Disclosure

MS COI Disclosure

SV COI Disclosure

Cover Letter

## Data Availability

Most of the data used in this study are freely available from the sources cited. The dataset is uploaded on Harvard DataVerse: Stephens, M., Lopez Berengueres, J.O., Moonesar, I.A. and Venkatapuran, S (2020), 1479 data points for COVID19 Policy responses, Harvard DataVerse. DOI:doi/10.7910/DVN/6VMRYG. The program is available by request to JB.

https://dataverse.harvard.edu/dataset.xhtml?persistentId=doi:10.7910/DVN/6VMRYG

## ACKNOWLEDGMENTS

The authors would like to thank the help we received Pavel Nesterov and Jorge Remon in modelling and Reem Gaafar for some data gathering.

## Contributors

This was a joint interdisciplinary project with each team member contributing to the project based on their expertise. MS conceived the study. MS and IAM acquired and managed the data, and IM conducted systematic reviews. JB analyzed the data and created the figures. MP, JP, and SV interpreted the results, and MS and IAM provided important local insights. MS, SV, and IAM drafted the first version of the manuscript with inputs from JB. The second version was redrafted by MS and SV with additional inputs from IM and JB. JB was responsible for visualization. MS supervised the work. MS is the guarantor and accepts full responsibility for the work and the conduct of the study, access to the data, and controlled the decision to publish. All co-authors provided critical comments and approved the final version of the manuscript. The corresponding author attests that all listed authors meet authorship criteria and that no others meeting the criteria have been omitted. MS has an innovation and strategy background. SV has a public health and ethics background. IAM has a global health policy background, and JB has a statistics and visualization background.

## Patient consent for publication

Not required.

## Ethical approval

Not required. All data sources used were fully publicly available. Further, the lead authors affirm that the manuscript is an honest, accurate, and transparent account of the study being reported; that no important aspects of the study have been omitted; and that any discrepancies from the study as initially planned (and, if relevant, registered) have been explained.

## Data sharing

Most of the data used in this study are freely available from the sources cited. The dataset is uploaded on Harvard DataVerse: Stephens, Melodena; Lopez Berengueres, Jose Oriol; Moonesar, Imanuel; Venkatapuram,Sridhar, 2020, “1479 data points of covid19 policy response times”, https://doi.org/10.7910/DVN/6VMRYG, Harvard Dataverse, V1, UNF:6:Uk0ndRTK8gKGJ01mKC960A== [fileUNF] The Kaggle code is available on request from JB.

## Dissemination to participants and related patient and public communities

Upon request, the data will be available for policymakers and government bodies.

## Patient and Public Involvment

Patients or the public WERE NOT involved in the design, or conduct, or reporting, or dissemination plans of our research

## Appendix 1: Spread of Disease for sample countries and key events (2019-2020)

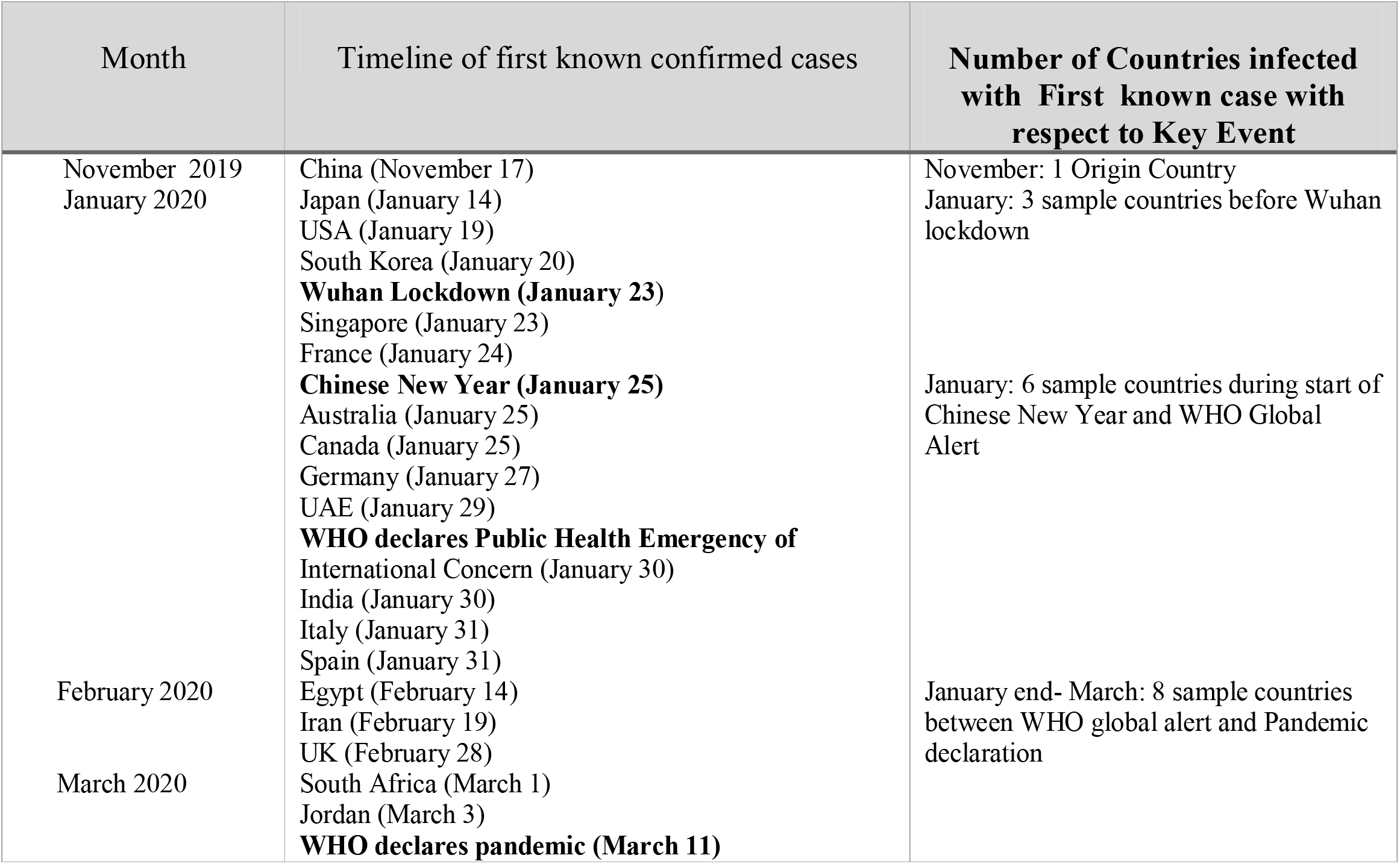

| Month | Timeline of first known confirmed cases | Number of Countries infected with First known case with respect to Key Event |
| --- | --- | --- |
| November 2019<br>January 2020 | China (November 17)<br>Japan (January 14)<br>USA (January 19)<br>South Korea (January 20)<br>**Wuhan Lockdown (January 23)**<br>Singapore (January 23)<br>France (January 24)<br>**Chinese New Year (January 25)**<br>Australia (January 25)<br>Canada (January 25)<br>Germany (January 27)<br>UAE (January 29)<br>**WHO declares Public Health Emergency of International Concern (January 30)**<br>India (January 30)<br>Italy (January 31)<br>Spain (January 31) | November: 1 Origin Country<br>January: 3 sample countries before Wuhan lockdown<br>January: 6 sample countries during start of Chinese New Year and WHO Global Alert |
| February 2020 | Egypt (February 14)<br>Iran (February 19)<br>UK (February 28) | January end- March: 8 sample countries between WHO global alert and Pandemic declaration |
| March 2020 | South Africa (March 1)<br>Jordan (March 3)<br>**WHO declares pandemic (March 11)** |  |

## Appendix 2: Systematic Review of Studies Using PRISMA 2009 Flow Diagram

### Keywords

COVID-19 ‘timing of policy’ or ‘policy timing’ or ‘timing of NPIs’ or ‘interventions’ or ‘policy interventions’; NPIs,’ or ‘nonpharmaceutical intervention’ or ‘non-pharmaceutical intervention’; ‘public health surveillance’; ‘epidemics health crisis’; ‘pandemics health crisis’; ‘outbreaks health crisis’; ‘outbreaks on mortality rates’; ‘time series analysis’; ‘health alert notice’; ‘workplace closure’; ‘border control’; ‘travel restriction’ or ‘travel precaution’; ‘school closure’; ‘case reporting’; ‘disinfection or decontamination’; ‘Infection control’; ‘public gathering’ or ‘group gathering’ or ‘group event’ or ‘public place’; ‘prevention’ or ‘mitigation’; ‘exit screening’ or ‘entry screening’ or ‘thermal screening’; ‘personal protective equipment’; ‘face mask’ or ‘facemask’ or ‘surgical mask’ or ‘N95’ or ‘respirator’; ‘hand hygiene’ or ‘hand washing’ or ‘handwashing’ or ‘hand disinfection’; ‘prevention’ or ‘mitigation’;

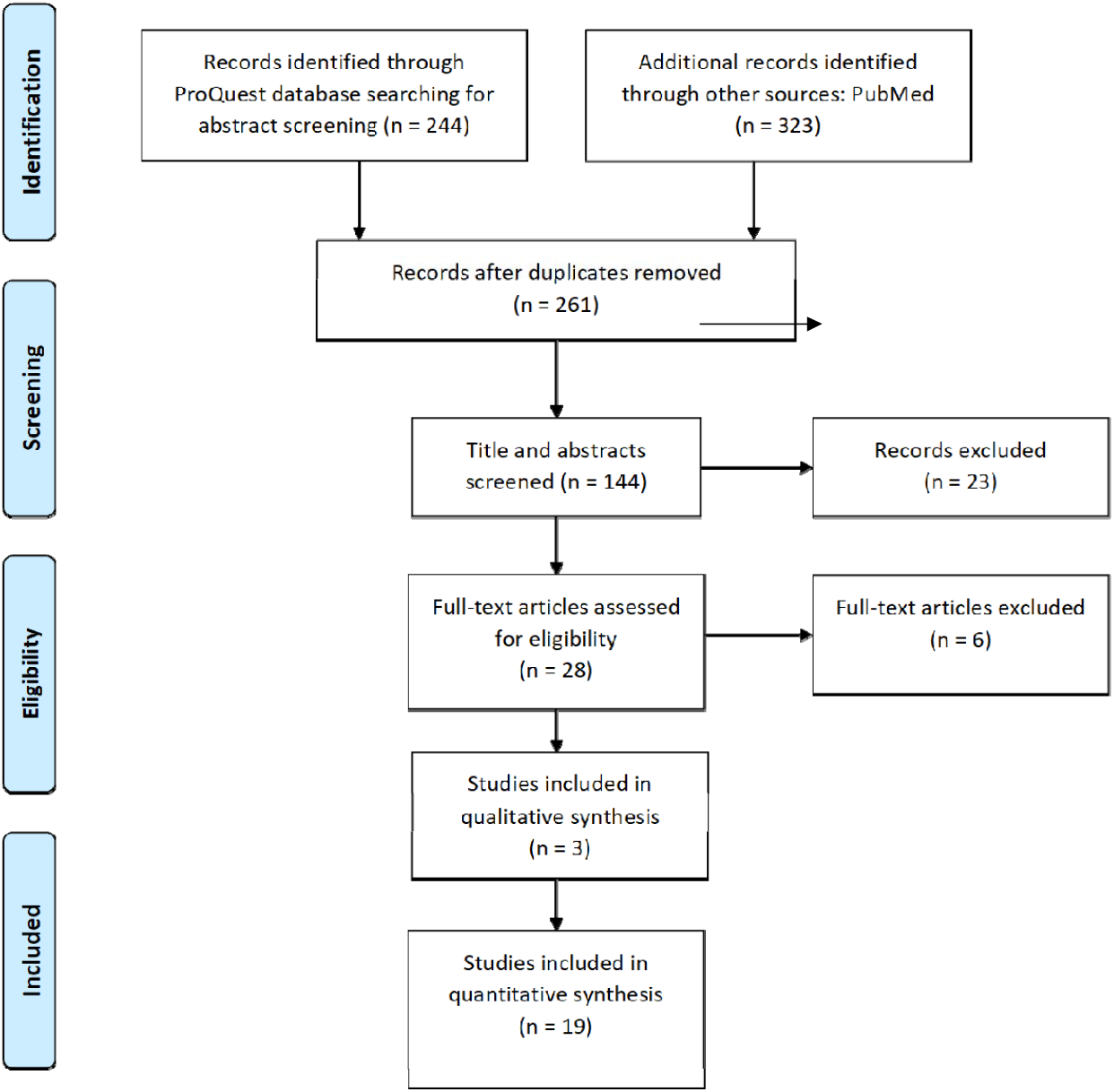

### Journals reviewed

PLoS One OR International Journal of Environmental Research and Public Health OR BMC Public Health OR PLoS Medicine OR BMJ Open OR PLoS Neglected Tropical Diseases OR Viruses OR Intensive Care Medicine Experimental OR Annals of Intensive Care OR PLoS Computational Biology OR PLoS Pathogens OR Scientific Reports (Nature Publisher Group) OR BMJ OR Clio Medica Online OR European Journal of Risk Regulation OR Globalization and Health OR Head & Neck OR BMC Infectious Diseases OR BMC Medicine OR BMJ Best Practice OR Epidemiology & Infection OR Global Health Action OR Homeland Security Affairs OR Infection Control & Hospital Epidemiology OR International Journal of Health Geographics OR Systematic Reviews OR An International Perspective on Disasters and Children’s Mental Health OR Asia & the Pacific Policy Studies OR BMC Health Services Research OR BMC Medical Research Methodology OR BMC Research Notes OR BMJ Global Health OR Clinical Medicine Insights. Oncology OR Disaster Medicine and Public Health Preparedness OR Health Expectations OR Health Research Policy and Systems OR Journal of Global Health OR Journal of Global Infectious Diseases OR Journal of Health, Population and Nutrition

