## Supplementary material for "Does the timing of government COVID-19 policy interventions matter? Policy analysis of an original database": Cover Letter

Melodena Stephens
Professor, Innovation Management
Mohammed Bin Rashid School of Government, Dubai, UAE


Editor
MedRxiv

15 September, 2020

**Sub:** New submission of an original research review article entitled, 'Does the timing of government COVID-19 policy interventions matter? Policy analysis of an original database."

Dear Editor-In-Chief,

I am pleased to submit an original research review article entitled, 'Does the timing of government COVID-19 policy interventions matter? Policy analysis of an original database' by Stephens, M., Berengueres, J., Venkatapuram, S., and Moonesar, I. A. for consideration as a preprint.

In this manuscript, the authors conducted a systematic review of the literature on policy responses and the timing of policy interventions, in addition to compiling an *original* database of COVID-19 policies (non-pharmaceutical interventions) across 17 countries. The present research studies the correlation between the timing of 87 COVID-19 policies across a group of 17 countries and their COVID-19 mortality rates. The data collection was conducted between April 6 2020 and June 4 2020. The data compiled covers the period between December 31 2019 to May 31 2020.

*What is already known in this topic?*
Recent scientific literature and public discussion have asserted that delays in implementing COVID-19 response policies resulted in thousands of preventable infections, and consequent deaths. These studies usually rely on dynamic mathematical models of infectious disease spread such as the well-known S-I-R model. Despite the public interest in government speediness in responding to the COVID-19 pandemic, we have found only 22 studies through a systematic review that examine some aspect of timing of interventions.

In terms of the breadth of policies, the most comprehensive database is the Oxford COVID-19 Government Response Tracker (OxCGRT). However, OxCGRT does not measure policy timings. Most studies that do focus on time, compare disease cases or the mortality rates before and after the time a policy is implemented. In this study, we are interested in measuring if implementing a policy earlier or later relative to three points in time (first Wuhan case, first national disease case, first national death) has any relation to the subsequent higher or lower mortality rates.

*What this study adds?*
We conduct a systematic review of literature in this topic and analyze an original database of 87 COVID-19 policy interventions in 17 countries from December 31, 2019 to May 31 2020. This study adds to existing literature on COVID-19 policies by evaluating the time-lag of policy implementation and their impact on mortality rates. The speediness of policy responses, in relation to three relevant points in time, do not by themselves have a correlation to mortality rates. Of the pandemic response policies that were implemented, there was no correlation between the time any particular policy was implemented and subsequent mortality rates. The findings also have implications for trans-national coordination of pandemic response policymaking. There is no consensus among countries on the appropriate timing from when an outbreak in a country is announced and when any particular policy should be implemented by other countries.

We hope you will consider our publication as it looks at the quickness of policy responses and its impact on the mortality rate of COVID-19. We have a five figures, three tables (three) and two appendices. We have also visualized our findings. The literature review is attached as a supplementary file as this is not the key focus of the paper and we hope to publish a separate paper on the same.

Our findings show that the timing of policy interventions across countries relative to the first Wuhan case, first national disease case, or first national death, is not found to be correlated with mortality. No correlation between country quickness of policy intervention and country mortality was found. Countries became quicker in implementing policies as time passed. However, no correlation between country quickness of policy intervention and country mortality was found. Policy interventions across countries relative to the first recorded case in each is not found to be correlated with mortality for 86 of the 87 policies. This suggests that there are other pre-existing contextual factors that need to be considered that are not highlighted by country rankings in reports like the Global Health Security Index. We find that having a higher score in the 2019 Global Health Security Index did not mean the country handled the pandemic better with respect to country-average delay in implementing a policy and country mortality. Hence our manuscript creates a paradigm for future studies on the timing of policy interventions.

This manuscript has not been published and is not under consideration for publication elsewhere. We have no conflicts of interest to disclose nor do we have objections to any reviewers for our manuscript.

I have also attached the source code for review if needed as a link in the appendix section and the details of the review as an appendix Excel spreadsheet attachment.

Thank you for your consideration.

Sincerely,

Professor - Innovation Management
Mohammed Bin Rashid School of Government
Level 7, Convention Tower, P.O. Box 72229, Dubai, UAE


Appendix

1. Source code : https://www.dropbox.com/sh/8i8n55j0e0jhl20/AAAdJ1kDTY3jkKp3LgdYPVE6a?dl=0

2. Review of 22 papers (attached as a supplementary file - Excel Sheet)
